# Development of a high-alert medicines list and pilot testing of error prevention strategies in Sri Lanka: a cross-sectional mixed methods study

**DOI:** 10.64898/2026.08.24.26361195

**Authors:** G.R.W.S.K. Bandara, N. Samaranayake, D. Ranaweera, D. Samaranayake, P. Galappatthy

## Abstract

High-alert medicines (HAM) cause preventable harm. This study aimed to develop a national HAM list and corresponding error prevention strategies (EPS) for hospital and community pharmacy settings in Sri Lanka. A mixed-methods cross-sectional observational study was conducted. Potential HAM were identified through an interviewer-administered questionnaire completed by 315 pharmacists from government (n=155) and private (n=160) sectors. Concurrently, a research pharmacist documented HAM-related problems and errors over three years. Additional HAM were identified by reviewing the Institute for Safe Medication Practices (ISMP) HAM list. A draft HAM list was refined and validated during five national stakeholder meetings convened by the Directorate of Healthcare Quality and Safety, Ministry of Health, Sri Lanka, involving 62 multidisciplinary stakeholders. These medicines were categorized using the A-PINCH (O) framework. EPS were developed through literature review and pilot-tested at a tertiary care hospital in Colombo. Key strategies included red-colour labelling, HAM stickers, separate storage, enhanced counselling, improved labelling, and patient information leaflets in Sinhala and Tamil for ten prioritised medicines. Feasibility was evaluated one year post-implementation using a questionnaire and three focus group discussions among hospital pharmacists (n=45). Quantitative data were analysed descriptively and qualitative data thematically. HAM identification differed by source: pharmacists (n=43), research pharmacist (n=40), ISMP list (n=80), and stakeholders (n=20). The validated list included 100 HAM for acute care settings, mainly injectable medicines, and 40 HAM for community settings, predominantly oral medicines. Awareness of the HAM list was 95.6% following implementation. Self-reported adherence to EPS, use of HAM labels and stickers, and availability of patient information leaflets were each 100%. Overall, 93.2% of pharmacists reported counselling patients on HAM requiring special instructions. Five themes emerged: usefulness, content adequacy, EPS implementation, operational barriers, and recommendations. National HAM lists for acute care and community settings were successfully developed. Pilot-tested EPS demonstrated high feasibility and acceptability, supporting national implementation and informing MOH policy through an official circular.

## BACKGROUND

High -alert medicines (HAM) can cause severe harm if used in error. Common examples include anticoagulants (e.g., warfarin), insulin, opioids, chemotherapeutic agents, and neuromuscular blockers, which are associated with increased potential for severe adverse outcomes such as bleeding, hypoglycaemia, respiratory depression, or death if errors occur [1–3]. These medicines are essential in healthcare, particularly in hospitals and specialized facilities, where ensuring their safe use is crucial.

Medication errors are preventable events that can arise at any stage of the medication use process, including prescribing, dispensing, administration, and monitoring [4,5]. Errors involving HAM are of particular concern because of these medicines’ narrow therapeutic indices and complex dosing requirements. Globally, HAM-related incidents have been identified as leading contributors to serious adverse drug events and patient harm in both hospital and community contexts [6]. Errors involving HAM can have severe consequences, including prolonged hospital stays, increased healthcare costs, and even death [7]. Hence, identifying HAM and implementing safeguards are essential components of patient safety.

Organizations must identify high-alert medicines (HAM) and implement safeguards such as standardized order sets, which are structured, evidence-based prescribing templates specifying the dose, route, frequency, and monitoring requirements to reduce variability and prevent errors [8].

Additional strategies include independent double-checks for high-risk orders, dedicated storage locations, and special labeling requirements to alert healthcare professionals and ensure safe administration[9,10]. Global frameworks such as those from the ISMP and the World Health Organization (WHO) High-Risk Medicines Initiative emphasize stringent safety measures [4,5,11–13]. The mnemonic “A-PINCHO,” recommended by the Australian Commission on Safety and Quality in Health Care and the World Health Organization in the Technical Report on Medication Safety in High-Risk Situations (2019), is used to identify groups of high-risk medicines [4,14,15].

Preventing HAM-related errors requires a multifaceted approach involving institutional policies, staff education, and technological support Countries and institutions are expected to have a HAM list for their local settings to be more careful during the use of these medicines [11,16–21]. The government and private sector pharmacies in Sri Lanka did not have a HAM list and private sector pharmacies in Sri Lanka did not have a HAM list, and pharmacists’ knowledge regarding HAM was minimal, according to a study conducted by the principal investigator [22,23].

In Sri Lanka, medication safety is challenged by system fragmentation between government and private sectors, limited resources, and a lack of standardized safety practices. Although there have been national efforts to improve patient safety infrastructure, there is no officially validated national HAM list or corresponding error prevention strategies (EPS) for pharmacy practice, according to previous studies conducted by the principal investigator, which highlighted gaps in HAM recognition, inconsistent counselling practices, and the absence of structured safety guidance for pharmacists [24,25]. This study aimed to address these gaps by developing a national HAM list with corresponding EPS and pilot testing them in a resource-limited setting to support safer use of HAM to prevent HAM-related errors across Sri Lankan pharmacy practice.

## METHODS

This observational cross-sectional study was conducted in three sequential phases using a mixed-methods approach that integrated quantitative surveys with qualitative stakeholder consultations and focus group discussions.

### Phase 1: Development of a draft high-alert medicines list (first draft) and error prevention strategies

A cross-sectional survey was conducted among 315 pharmacists from government (n = 155) and private (n = 160) sectors. All government pharmacists working in all outpatient and inpatient pharmacies and drug stores at two tertiary-care teaching hospitals in the Colombo District, Sri Lanka, Hospital 1 (H-1; 3,318 beds) and Hospital 2 (H-2; 1,279 beds) were recruited. In addition, eighty private pharmacies located within a 3-km radius of the two hospitals were included according to International Network for Rational Use of Drugs (INRUD) guidelines to identify high-alert medicines and assess medication-safety practices [26].

Data were collected using an interviewer-administered questionnaire (IAQ) developed in-house to assess medication-safety practices related to high-alert medicines (HAMs) among participating pharmacists. The IAQ was content-validated by two pharmacy experts, while face validity was assessed by five pharmacists from another hospital who were not involved in the study. Data collection was conducted at H-1 from 21.04.2018 to 23.01.2019, at H-2 from 25.01.2019 to 20.01.2020, and at private pharmacies from 23.01.2020 to 10.10.2021. The data-collection period was interrupted by the COVID-19 pandemic.

Following written informed consent, participants completed the IAQ, which assessed their knowledge and awareness of HAMs. They were also asked to identify medicines they had encountered in their practice that had resulted in severe patient harm or near-miss incidents in the local setting. The medicines identified by participants were collated to develop a preliminary HAM list relevant to the Sri Lankan context. This list was subsequently supplemented with medicines included in the Institute for Safe Medication Practices (ISMP) HAM list and additional medicines were identified by the principal investigator (study pharmacist) based on medication-related problems encountered over a three-year period, from 25.01.2019 to 23.01.2022, and on professional experience.

Error prevention strategies (EPS) specifically targeting HAMs were developed by the principal investigator based on HAM-related problems encountered during the study, professional experience, and evidence from the published literature.

### Phase 2: Stakeholder consultation and development of recommendations

Five national-level consultative meetings were held from 17.08.2023 to 26.09.2023 at the Directorate of Healthcare Quality and Safety (DHQS) of the Ministry of Health. Sixty-two stakeholders participated, including clinical pharmacologists, physicians, paediatricians, anaesthetists, psychiatrists, administrators, pharmacists, and nurses representing both government and private sectors.

The first draft of the HAM list based on Phase 1 studies was presented during the five consultative meetings for content validation. Feedback, including divergent opinions and practical examples was incorporated to finalize the HAM list and vigilant points noted by experts at consultative meetings were included to the HAM list as a column. Revised versions of the list and the proposed error prevention strategies (EPS) were circulated among the meeting participants for additional feedback, which was then integrated to finalize the EPS.

#### Preparation of the HAM list

Responses of stakeholders at consultative meetings were aggregated, duplicates were removed, and medicines were categorised by severity and medicine class using the A-PINCH-(O) mnemonic. Separate HAM lists were prepared for acute-care and community settings. Specific vigilance points were added to improve clarity and clinical applicability.

#### Implementing of suggested error-prevention strategies on HAM

The error prevention strategies (EPS) for HAM were developed by the principal investigator during Phase 1, following a comprehensive review of the literature on preventing HAM-related errors, and were finalized in Phase 2. Main EPS recommended were labelling as ‘HAM’ in red colour, using HAM stickers, HAM warning labels and storing HAM separately. EPS included independent double-checking during preparation, dispensing, and administration, supported by adherence to the “five rights” of medication safety. Patient-centred measures, such as counselling and provision of written instructions, were incorporated to promote safe use beyond hospital settings. The strategies also emphasized continuous monitoring, accurate documentation, non-punitive error reporting, and regular staff training.

A selected list of ten HAM; warfarin, methotrexate, alendronate, insulin, lithium carbonate, azathioprine, sodium valproate, oral morphine, erythropoietin, and digoxin; was prioritized for focused interventions based on locally reported errors and incidents. Patient information leaflets (PIL) were developed for these medicines in English, content validated by experts, and was translated to two main local languages (Sinhala and Tamil). These selected HAM were the focus of targeted patient counselling, provision of PIL, and the development of specific labels to further enhance safe use.

### Phase 3: Pilot testing of HAM list and error prevention strategies and feasibility assessment

The final HAM list was nationally approved and circulated as an official circular (Circular No. 01-39/2024) by the Ministry of Health [27]. It was distributed to all healthcare institutions and hospitals in Sri Lanka through the heads of institutions. District (Hospital H-2) was purposively.

For pilot testing, a tertiary-care hospital in Colombo selected to assess the applicability of the national HAM list in routine clinical practice and the feasibility of implementing the recommended EPS. The hospital was selected as a representative tertiary-care setting with multiple pharmacy units, wards, and other clinical areas where HAM medicines are prescribed, dispensed, stored, and administered. The HAM list and EPS were introduced across relevant pharmacy units, wards, and other hospital units involved in the medication-use process.

Pilot testing of the above-mentioned EPS for HAM was conducted in all outpatient and inpatient pharmacies, wards and clinical units of Hospital H-2. The intervention involved disseminating the national HAM list and EPS to pharmacies, wards and clinical units through Quality Management Unit (QMU) of the hospital.

Before implementation of these EPS, These medicines were not stored separately and no warning signs were used to differentiate them from non-HAM in pharmacies, wards, and clinical units. According to the EPS, red HAM warning stickers and auxiliary warning labels were affixed to identify HAM, and storage arrangements were reorganized according to pharmacological class (Fig 1).

**Fig 1.**
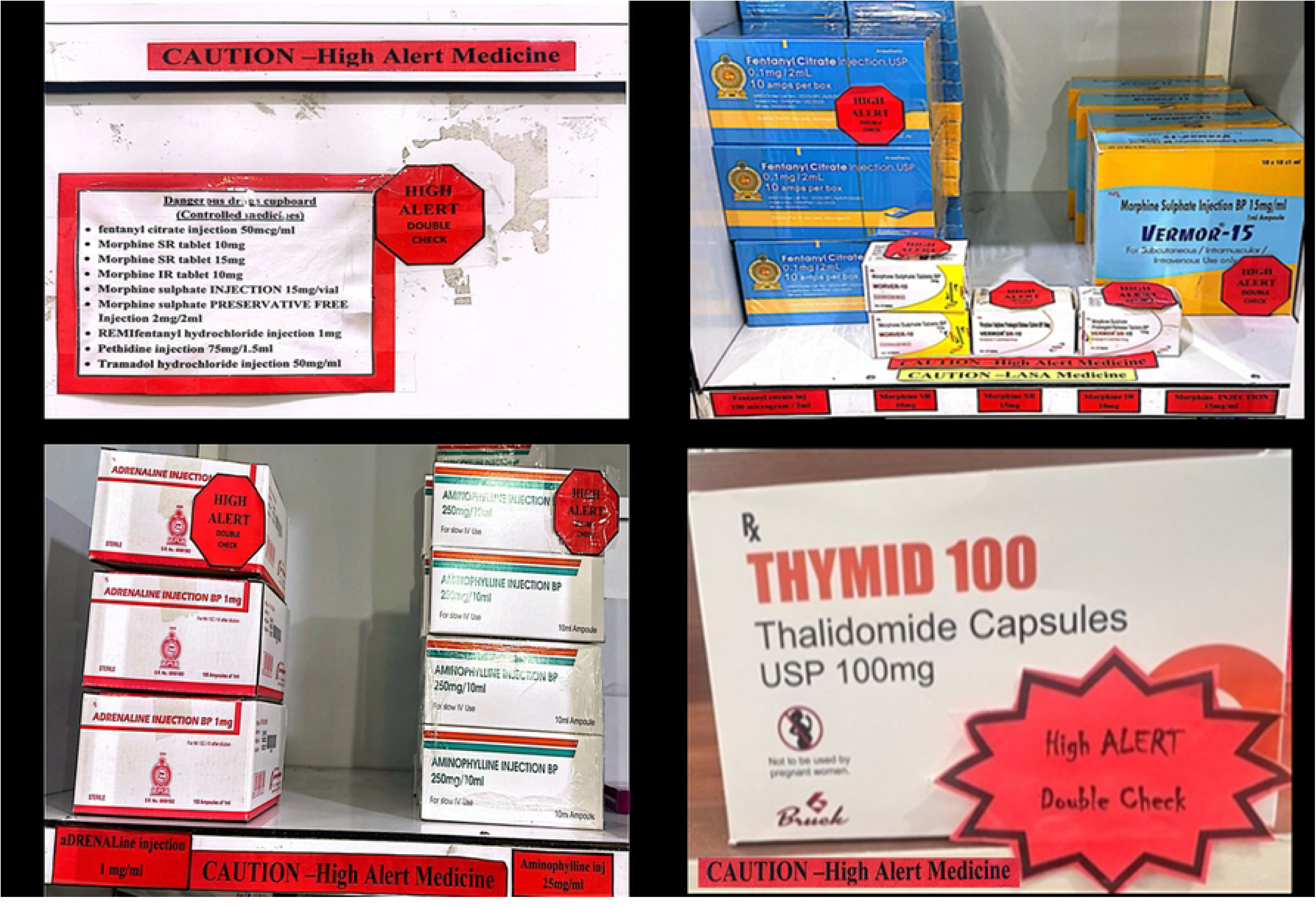
High-alert medication storage and labeling interventions implemented at in-patient pharmacy in Hospital H-2 during pilot testing. (A) Controlled medicines cupboard labelled with “CAUTION - High alert medicine,” “HIGH ALERT“-Double check” sticker and the list of stored narcotics. (B) Fentanyl, morphine injections and morphine tablets storage with red “HIGH ALERT” stickers applied as part of error prevention strategies. (C) Adrenaline and aminophylline injections with shelf-edge “CAUTION - HIGH ALERT MEDICINE” labels and stickers designed for this study. (D) Storing thalidomide capsules with “High ALERT -Double check” starburst sticker and warning labels. All labels and stickers were developed and placed by the principal investigator for visual identification of HAM. Photographs taken with permission, no patient-identifiable information is shown.

Targeted safety checks were introduced at prescribing, dispensing, and administration stages. Counselling clinic-patients at first dispensing was carried out and written instructions were provided in local languages, to promote safe use at home (Fig 2).

**Fig 2.**
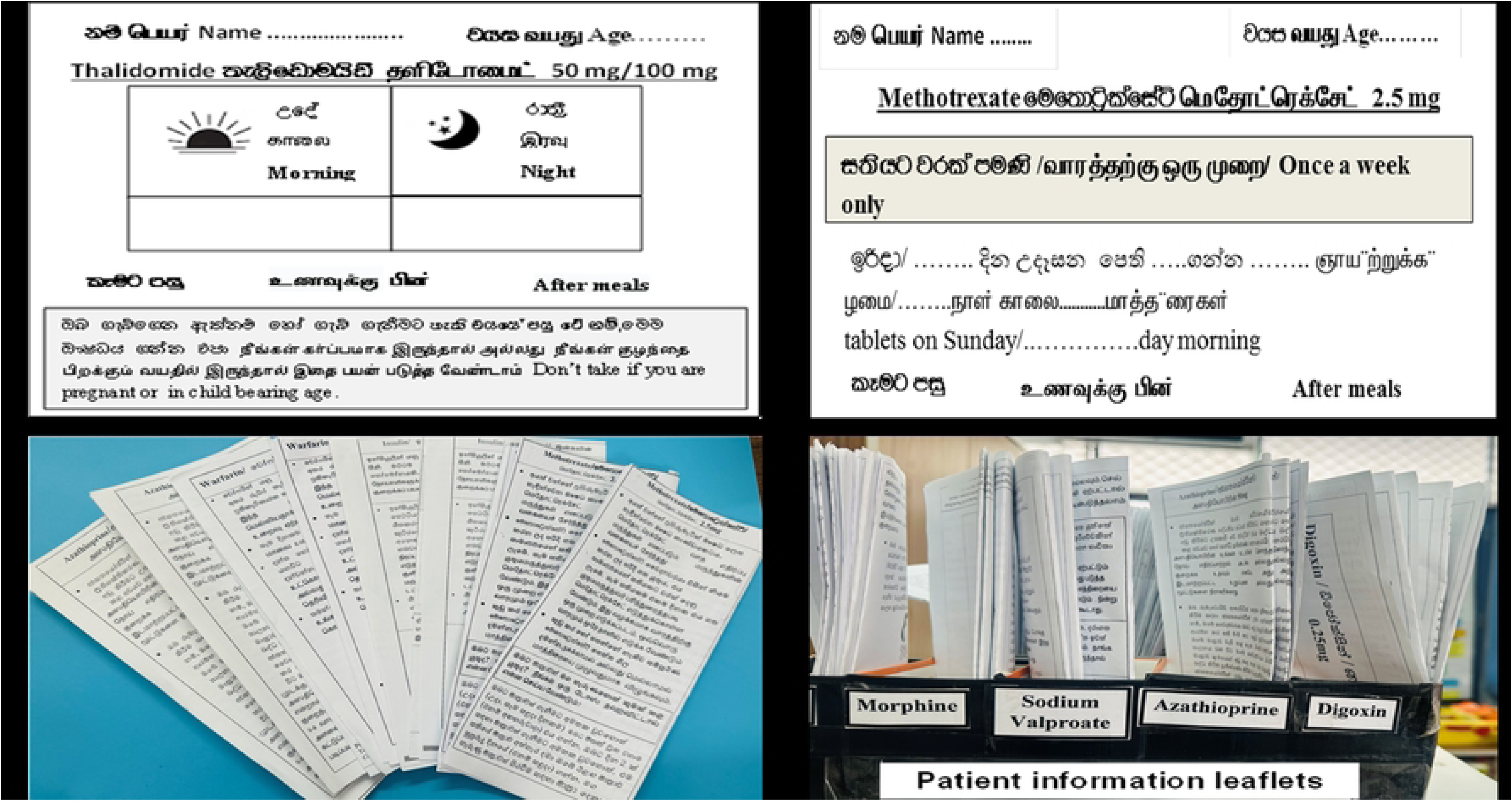
Tri-lingual dispensing labels and patient information leaflets with dosing instructions for high-alert medications, at outpatient (clinic) pharmacy in Hospital H-2 for medication safety in Sri Lanka. (A) Sample thalidomide dispensing label showing dosage, timing, and teratogenicity warning in Sinhala, Tamil, and English. (B) Sample methotrexate leaflet with “Once a week only” instruction highlighted in Sinhala, Tamil, and English. (C) Examples of set of patient information leaflets developed for different high-alert medicines in local languages, identified in the study. (D) Storage system for printed leaflets labelled by medicine name: morphine, sodium valproate, azathioprine, and digoxin at dispensing counters in outpatient (clinic) pharmacy. Leaflets were designed by the principal investigator to improve patient safety and understanding of high-risk medications. Photographs of prototype materials taken with permission. No patient-identifiable information is shown.

Pharmacists provided patient information leaflets (PIL) during counselling to educate patients on the safe use of HAM.

The pilot enabled assessment of feasibility, staff compliance, and operational challenges and generating practical insights to refine the strategies for subsequent hospital-wide implementation.

Feasibility was evaluated one year after pilot testing using a structured interviewer administered questionnaire (IAQ), developed in house and three focus group discussions (FGDs) conducted among all pharmacists at H-2 (n = 45) from 30.06.2024 to 30.08.2024. The IAQ evaluated HAM list and EPS implementation, including pharmacists’ perceptions of challenges and adherence. Content validity was assessed by two medication safety experts, and face validity by seven independent pharmacists not involved in the study. For the FGDs, a co-facilitator served as note-taker and research supervisors observed the sessions. Informed written consent was obtained from all participants for both quantitative and qualitative studies. Written and verbal invitations informed participants about study objectives, procedures, and confidentiality. Discussions were conducted until thematic saturation was reached; each group included approximately 15 pharmacists and lasted about 2.5 hours. A semi-structured guide aligned with the HAM list and EPS explored perceptions, implementation experiences, barriers, and recommendations in the FGDs. Sessions were audio-recorded and supplemented with field notes.

The first two FGDs were attended by supervisors to support facilitation and to refine the process; the final FGD was led by the principal investigator. Discussions were conducted in Sinhala, translated into English, and transcribed verbatim. Transcripts were cross-checked against audio recordings and field notes for accuracy.

Quantitative data were analysed using descriptive statistics in SPSS version 21. Qualitative data were analysed using a hybrid deductive–inductive thematic approach guided by Braun and Clarke’s six-step thematic analysis approach [25,28]. An initial deductive coding structure was developed from the discussion guide, while inductive codes were generated from the data and grouped into subthemes through constant comparison. Representative quotations were selected to illustrate each theme and enhance transparency.

#### Ethics approval

Ethics approval and consent to participate was approved by the Ethics Review Committee of Faculty of Medicine, University of Colombo (Reference number: EC-18-008) and Ethical Review Committees of National Hospital of Sri Lanka (NHSL) (Reference number: AAj/ETH/COM/2017) and Colombo South Teaching Hospital (CSTH) (Reference number: PL/MO/2018-2019). Approval was renewed annually and covered the full study period. Permissions from all selected community pharmacies were obtained prior to commencing the study. Participation was voluntary, and informed written consent was obtained from all participants. Administrative permission to photograph medication storage areas was obtained from the Chief Pharmacist/In-charge of Pharmacy at hospital. All photographs were de-identified and contained no patient-identifiable information or images of people.

## RESULTS

### Identification and Validation of HAM

Pharmacists initially identified 43 high-alert medicines (HAM), which were supplemented by 80 medicines from the Institute for Safe Medication Practices (ISMP) list and a further 40 proposed by the research pharmacist. During stakeholder consultations, 20 additional HAM were suggested. From an initial pool of 163 medicines, 100 HAM were shortlisted and finalised at the consultative meetings [S1 Table. Total list of HAM for community settings (total=40)].

The final HAM list comprised 100 medicines for acute-care settings and 40 for community settings, according to Table 1.

**Table1.** Distribution of high-alert medicines by dosage form in acute care and community settings, Sri Lanka.

| Types of dosage forms | Acute care settings<br>(n = 100) |  | Community settings<br>(n = 40) |  |
| --- | --- | --- | --- | --- |
|  | n | % | n | % |
| Oral preparations | 30 | 30.0 | 36 | 90.0 |
| IV/ injectable preparations | 62 | 62.0 | 4 | 10.0 |
| Both oral and injectable preparations<br>(E.g. oral and injectable amiodarone) | 8 | 8.0 | - | - |
| Total | 100 | 100.0 | 40 | 100.0 |

In acute-care settings, including hospitals and intensive care units (ICUs), injectable and intravenous formulations predominated, accounting for 62% of the list, according to Table 2. In community settings; such as community pharmacies, outpatient departments, and dispensaries; 40 HAM were identified, of which 90% were oral formulations (S2 Table. Total list of HAM for community settings -total=40).

**Table 2.**
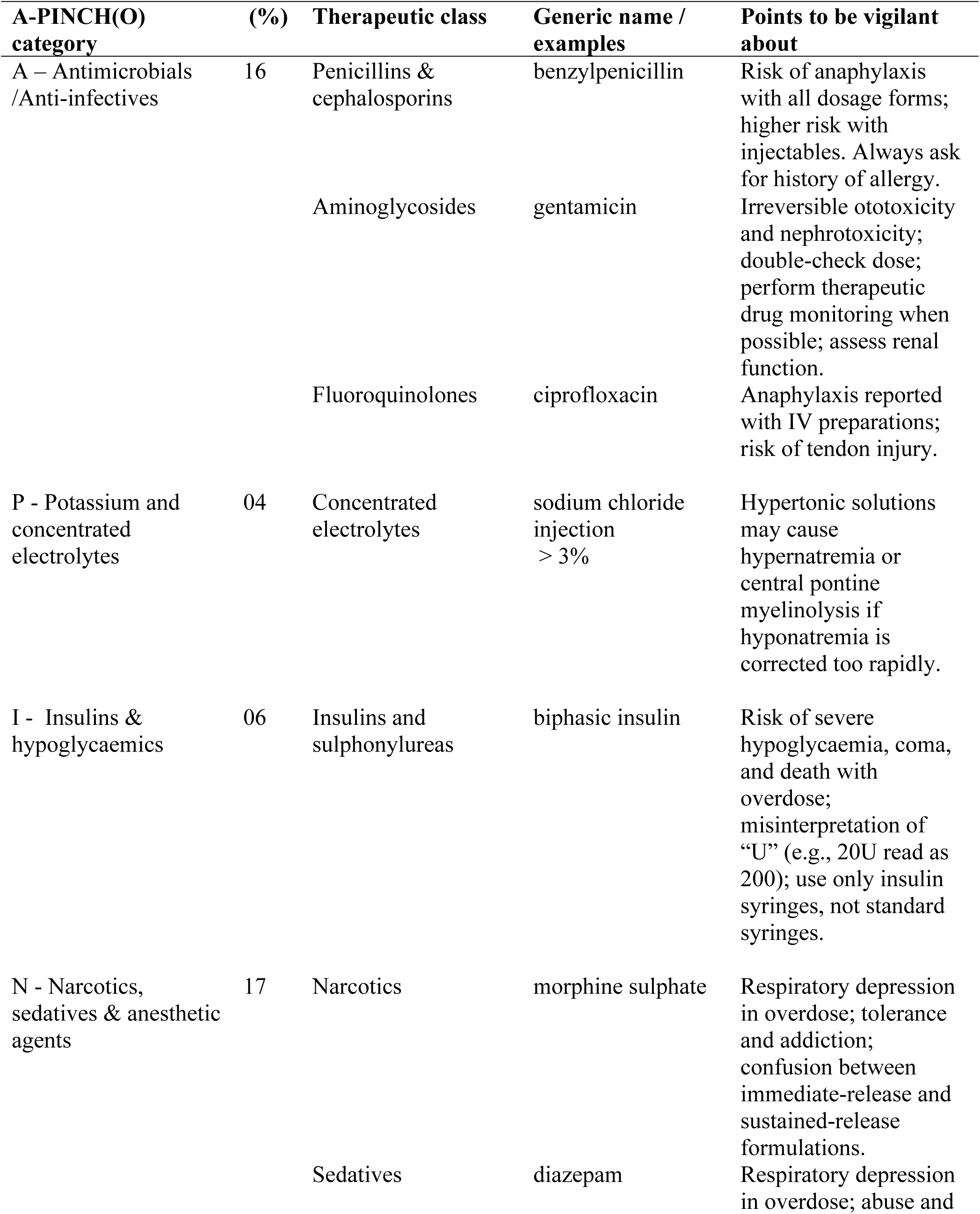
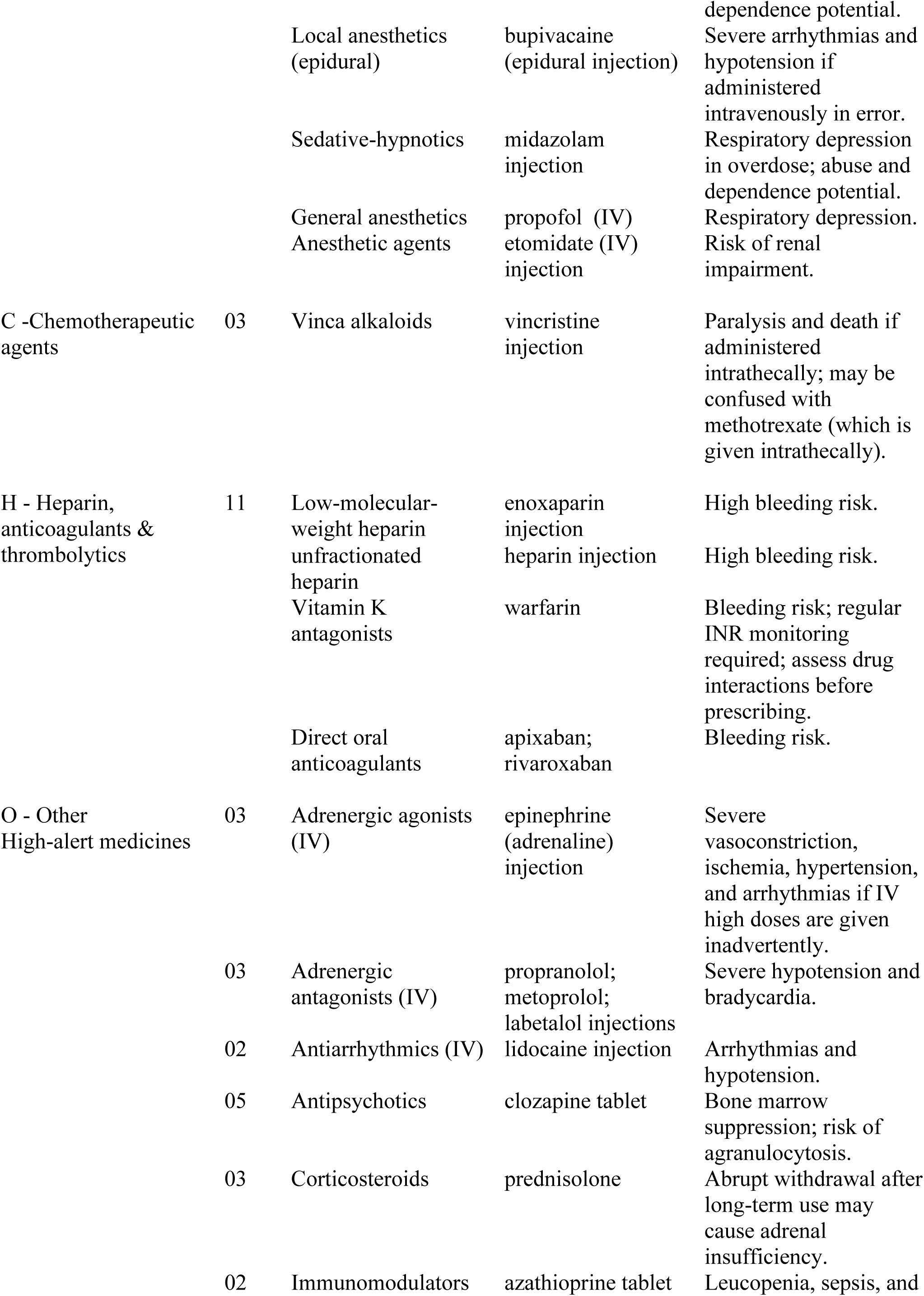
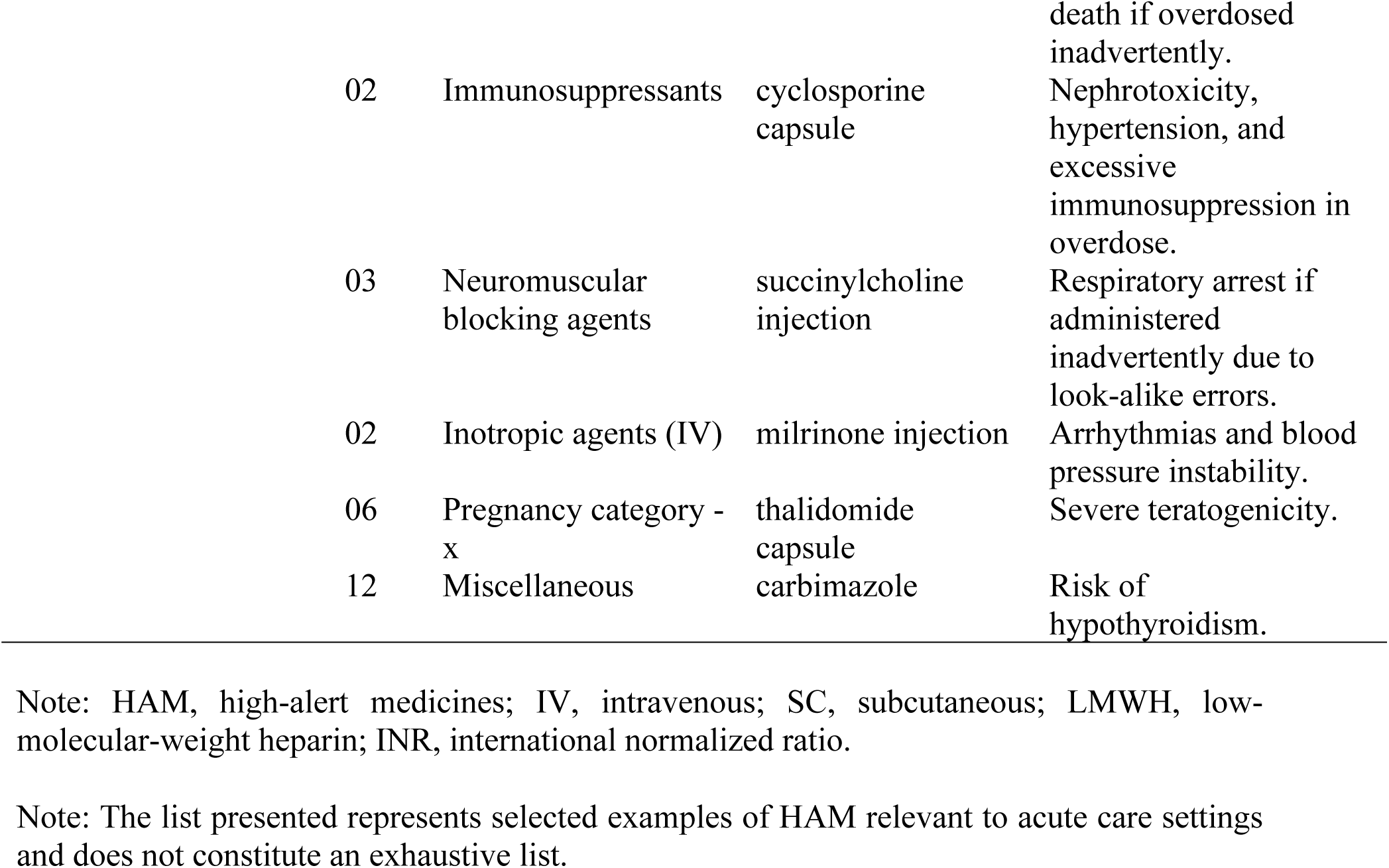
High-alert medicines by A-PINCH (O) category with examples and safety points in acute care settings, Sri Lanka (n = 100)

Table 1 presents the distribution of HAM by type of dosage forms in acute-care and community settings.

### Categorization of HAM

Categorization using the APINCHO framework showed that the largest proportion of HAM fell under the “Other medicines” category (43%), which included medicine such as adrenergic agonists and antagonists, antiarrhythmics, corticosteroids, and neuromuscular blocking agents. A relatively high proportion of narcotics, sedatives, and anaesthetic agents (17%) were also included in the list, Table 2 summarizes the distribution of HAM by APINCHO category.

### Feasibility Assessment of HAM Error Prevention Strategies

#### Quantitative study

##### Characteristics of participants

A total of 45 pharmacists from outpatient, inpatient pharmacies and stores participated, comprising 11 males (24.4%) and 34 females (75.6%). The majority were aged 33–44 years (51.1%), and 28.9% had more than 20 years of work experience. Regarding educational qualifications, 35.6% held a B.Pharm or B.Sc. Pharmacy degree, 55.6% had a Diploma or Higher Diploma in Pharmacy, and 8.9% possessed a degree in another discipline. In terms of workload, 28.9% reported dispensing 200–300 prescriptions per day, while 24.4% dispensed 300–400 prescriptions per day.

Pharmacists’ perspectives on high-alert medications (HAM), safe practices, and perceived barriers to implementation, obtained from the quantitative study, are summarized in Table 3. The reported barriers to providing patient counselling on HAM, obtained from the quantitative study, are shown in Table 4.

**Table 3.**
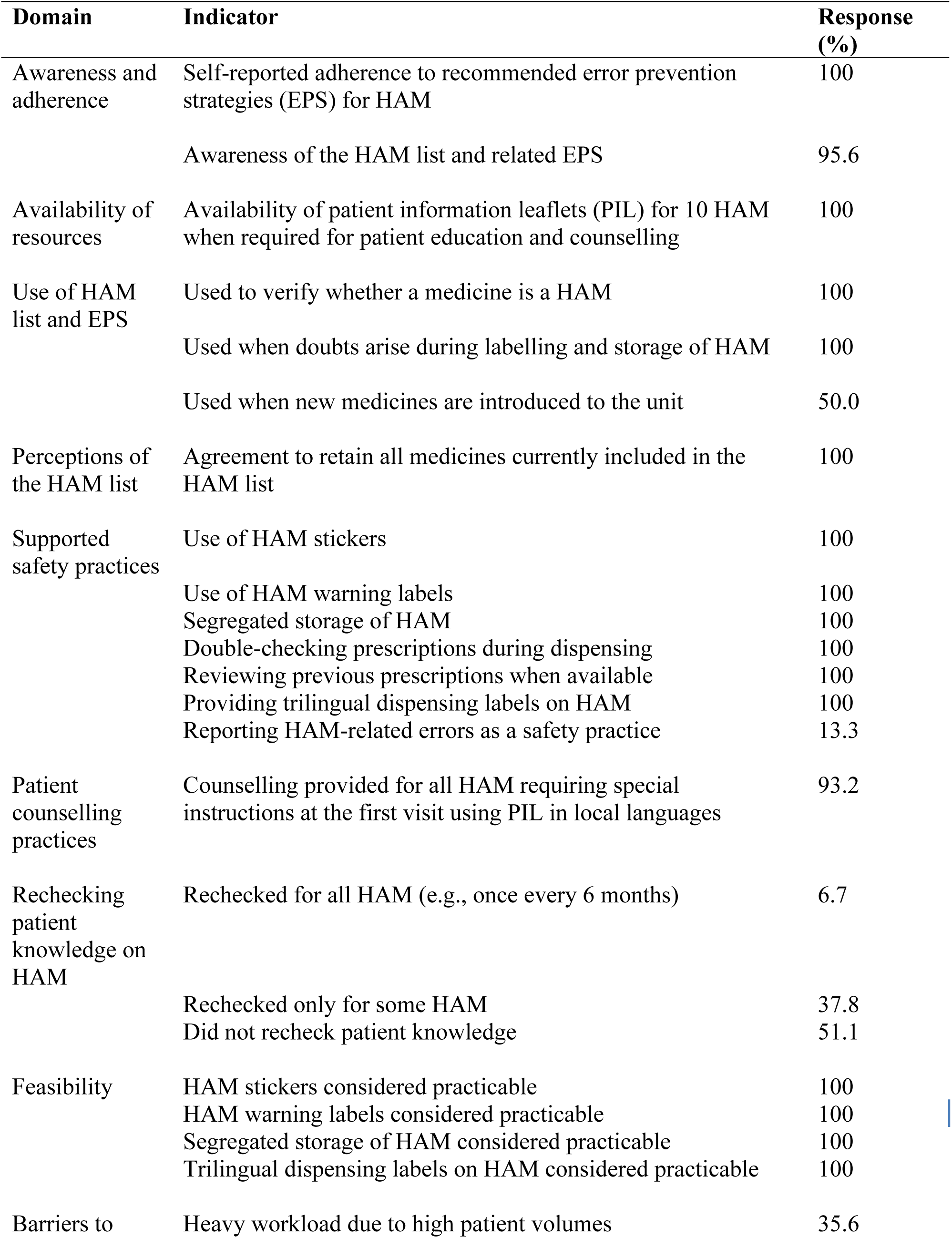
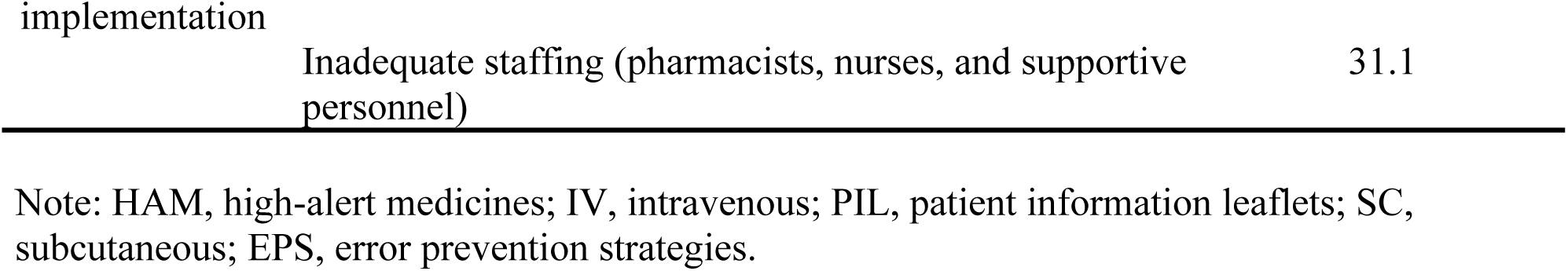
Pharmacists’ perspectives, practices, and barriers related to high-alert medicines (n = 45), Sri Lanka.

**Table 4.** Reported challenges to providing patient counselling on high-alert medicines (n = 45), Sri Lanka.

| Category of challenges | Specific challenge | % of respondents |
| --- | --- | --- |
| Patient-related barriers | Resistance to learning about medicines | 24.4 |
|  | Lack of motivation to learn | 26.7 |
|  | Language barriers | 2.2 |
|  | Differing health literacy levels | 37.8 |
|  | Combination of all above factors | 8.9 |
| Pharmacist-related barriers | Fatigue due to heavy workload | 35.6 |
|  | Time constraints due to long patient queues | 35.6 |
|  | Inadequate pharmacy staffing | 26.7 |
|  | Combination of all above factors | 2.2 |
| Technological / system-related barriers | Limited access to reference resources (e.g. no Wi-Fi, BNF) | 8.9 |
|  | Absence of a designated patient counselling area | 91.1 |

Note: BNF, British National Formulary.

#### Qualitative study

As part of the qualitative component of the study assessing medication safety practices and associated error prevention strategies, including HAM-related practices, three focus group discussions (FGDs) were conducted with all 45 hospital pharmacists who participated in the pilot implementation. Five major themes were identified from three FGDs; perceived usefulness of the HAM list, comprehensiveness and adequacy of the HAM list, implementation of EPS, operational barriers to EPS implementation, and suggestions for strengthening EPS.

##### Theme 1: Perceived usefulness of the HAM list

Across all FGDs, pharmacists unanimously acknowledged the availability and utility of the HAM list within their work units. They described it as an essential reference tool that is used daily, particularly during dispensing, issuing medication to wards, and when uncertainty arises.

Participants emphasized that the list significantly contributes to reducing medication errors by highlighting medicines that require extra vigilance.

Supporting quotes:

> “HAM list is very useful; when we issue medications to wards and units, we double check HAM before issuing.” **–** P2, FGD-1
>
> “We normally use it every day and also when a doubt occurs.” – P3, FGD-3

##### Theme 2: Adequacy and comprehensiveness of the HAM list

The list was generally considered comprehensive, pharmacists agreed that the HAM list used during pilot testing was adequately detailed, and required no additions. The list was consistently described as thorough and clear, supporting its use in routine practice. Although the exclusion of frequently used intravenous (IV) antibiotics was initially proposed, this suggestion was later rejected. Participants agreed that these medicines should be retained due to their association with adverse drug reactions and the risk of serious harm when administered incorrectly.

As noted by participants, “The HAM list is descriptive enough. The potential risks are included” (P1,( Chief Pharmacist), FGD-3), and “We initially thought of suggesting to remove IV antibiotics to reduce the list, but due to serious reactions such as anaphylaxis experienced, we are agreeable to keep them” (P4, FGD-1).

##### Theme 3: Implementation of HAM error prevention strategies (EPS)

Adherence to the recommended error prevention strategies (EPS) for HAM was reported to be acceptable across all practice settings. Separate storage of HAM was universally implemented, with medicines kept in designated cupboards or specific refrigerator shelves and supported by visible HAM lists for quick reference. Visual risk communication using red warning labels and HAM stickers was routinely practiced and perceived as effective in promoting vigilance during dispensing.

Double-checking procedures were commonly applied, particularly for prescriptions containing multiple HAM and during high-volume outpatient dispensing, where limited time increased the risk of error. Reviewing previous prescriptions was also a routine practice, especially in clinic settings, to prevent duplication and LASA-HAM-related errors. For IV injectable antibiotics, indoor pharmacy staff consistently used auxiliary labels or printed instructions to guide correct preparation and administration by nursing staff.

Illustrative comments included: “We use a separate cupboard and upper shelves in refrigerators for HAM medicines” (P5, FGD-1), “Red HAM labels are eye-catching to be vigilant when we issue them*”* (P6, FGD-2), and “Some prescriptions contain more than five HAM… double-checking is useful because we have only about 30 seconds per patient” (P8, FGD-1).

##### Theme 4: Barriers and operational constraints

Despite strong adherence to EPS, pharmacists identified several operational challenges that limited optimal implementation. Lack of physical space restricted separate storage of HAM in some pharmacies, while inadequate infrastructure-including shortages of computers, Wi-Fi access, and colour printers affected labelling and documentation practices. High workload and overcrowding, particularly in outpatient departments, resulted in extremely short dispensing times and increased pressure on pharmacists.

Participants also raised concerns about nursing practices, noting that administration instructions and manufacturer leaflets for IV antibiotics were often overlooked, increasing the risk of administration-related errors. One participant explained, “HAM stickers would be clearer if printed in red on white background, but we don’t have colour printers” (P4, FGD-2), while another stated, “Nurses normally don’t read the manufacturer’s leaflet… slow administration is essential for some IV antibiotics” (P5, FGD-1).

##### Theme 5: Suggestions for strengthening the EPS

Participants recommended several strategies to further strengthen HAM-related safety practices. Key suggestions included improving guidance on IV antibiotic administration, particularly emphasizing slow infusion using burette sets for medicines such as co-amoxiclav and ciprofloxacin. Strengthening structured education and regular training for nursing staff was also highlighted as essential to reduce administration-related errors.

While some pharmacists suggested a multi-colour coding system to indicate different levels of risk, most agreed that maintaining a single red warning colour was more practical and easily recognisable in busy clinical settings. Additional recommendations focused on improving infrastructure, including increasing storage space and providing essential technological resources to support effective EPS implementation. As summarized by one participant, “Maybe multiple colours… but finally red is ok for easy recognition” (P9, FGD-3).

## DISCUSSION

This study represents the first comprehensive national effort in Sri Lanka to identify HAM and implement structured EPS for HAM related errors in both outpatient and inpatient settings. It is also probably the first published study from a low and middle income country (LMIC) developing and pilot-testing a HAM list in a real-world hospital environment, generating evidence using mixed methodology that led to the issuance of a MOH circular on HAM and EPS, marking a major step forward in medication safety governance [27].

The study produced the first standardized HAM list for Sri Lanka, combining international ISMP classifications with local expert input. The final list, comprising 100 medicines for acute settings and 40 for community settings, reflects global patterns in HAM categorization. The predominance of injectable preparations in acute settings aligns with international evidence indicating that intravenous medicines pose a significantly higher risk of severe harm due to narrow therapeutic windows and complex administration procedures [4,5,13,16]. Categorization using the APINCHO framework revealed that the “Other medicines” group accounted for 43%, consistent with international HAM lists in which adrenergic agonists, neuromuscular blockers, and antipsychotics are frequently included due to their risk profile [13,16]. Ten medicines with recurrent local errors, such as insulin and warfarin, were prioritized, paralleling global findings on high-risk outpatient medicines that commonly cause preventable hospitalizations [27].

The development of targeted EPS is arguably as critical as the HAM identification itself. Strategies proposed for acute care settings address the key risk points and well-supported by international evidence as reducing errors when robustly implemented [4,11,13,29,30].

Quantitative results showed total adherence to main EPS components, including the use of HAM stickers, segregated storage, reviewing previous prescriptions. Pharmacists attended to independent double-checks for HAM, dedicated storage with auxiliary labels, structured patient counselling using teach-back methods, prescription reviewing for drug interactions and contraindications, screening patient medication records, and proactive monitoring systems (e.g. INR for warfarin). Qualitative findings supported this, with pharmacists describing the strategies as essential, highly visible, and crucial during high-volume dispensing workflows. These findings mirror studies from the USA, Canada, and Australia, where strong EPS for HAM uptake has been associated with improved medication safety culture and reduced incidence of errors [30–34]. In Sri Lanka, this level of compliance is particularly noteworthy given the constraints of a resource-limited public-sector healthcare environment.

Pilot testing demonstrated that HAM-related safety practices were feasible and well accepted, even under high-pressure conditions. The list was reported to be comprehensive, and the safety strategies were widely implemented across units. Pharmacists displayed high awareness and proactive engagement in medication safety activities. Pharmacists not only followed EPS but also adapted them creatively, attaching auxiliary administration labels for intravenous co-amoxiclav, reorganizing storage according to drug class, and visually marking HAM cupboards. These practices reflect a proactive safety culture comparable to international hospitals where standardized HAM labelling and storage protocols have been integrated into daily workflows [11,30,33,34].

Despite high adherence, pharmacists consistently highlighted barriers including heavy workload, staff shortages, inadequate space, and limited access to digital tools. Over one-third of participants (35.6%) reported time pressure and fatigue, while 91.1% indicated that the absence of a dedicated counselling area limited optimal service delivery. These challenges are consistent with findings from LMIC, where insufficient infrastructure and human resources frequently impede medication safety implementation [35]. A notable systems-level gap identified was the inconsistent reading of manufacturer instructions by nurses, especially for intravenous antibiotics requiring slow infusion. Similar administration-related errors linked to inadequate training and high workload have been reported in studies from South Asia, including Nepal and India [36].

Although most pharmacists (93.2%) educated first-visit patients on HAM, only 6.7% periodically reassessed patient knowledge. Qualitative data indicated that poor health literacy, low motivation, and resistance to counselling hindered effective patient engagement, a challenge echoed in global studies on high-risk outpatient medicines in resource-limited settings [37]. Limited access to Wi-Fi and reference tools, reported by 8.9% of participants, further restricted the ability to provide up-to-date medication information for counselling.

Sri Lanka’s EPS, including segregated storage, visual warning cues, and double-checking procedures, closely follow recommendations from the WHO and the ISMP for mitigating risks associated with HAM. The high uptake demonstrates that structured safety strategies can be successfully implemented even in busy tertiary care hospital settings with minimal resources. However, unlike high-income countries that utilize electronic alerts, barcode scanning systems, and automated dispensing technologies, medication safety in Sri Lanka relies heavily on manual processes, increasing the risk of human error. Although pharmacists suggested more advanced color-coding systems, a consensus was reached to retain a single red label to minimize confusion, an approach supported by research warning against the risks of “over-color-coding” in medication safety systems [37,38].

This study promotes standardization of safety practices across the healthcare system, reducing errors and facilitating consistent training and quality improvement initiatives. Relying on institution-specific lists may lead to significant variations and weaken national safety efforts [4].

A major strength of this study is its direct policy impact. The validation process involved 62 stakeholders, including six professors, eight specialists representing national professional colleges (The Sri Lanka College of Clinical Pharmacologists and Therapeutics, Ceylon College of Physicians, Sri Lanka College of Psychiatrists, Sri Lanka College of Anesthesiologists, Sri Lanka College of Surgeons, Sri Lanka College of Obstetricians and Gynecologists, Sri Lanka College of Internal Medicines and Sri Lanka College of Pediatricians), 16 medical officers, seven administrators, ten pharmacists, seven nursing officers, and eight healthcare professionals from the private sector. This collaborative engagement resulted in the issuance of a formal circular by the Ministry of Health (MOH) endorsing the HAM list and associated safety strategies [27]. The use of a mixed-methods approach further strengthened the validity of the findings, as quantitative data on adherence were triangulated with qualitative insights related to feasibility, perceptions, and implementation challenges. In addition, real-world pilot testing conducted across multiple hospital units, including outpatient departments, clinics, and wards, demonstrated practical feasibility while identifying operational barriers critical for planning nationwide scale-up. Strong staff acceptance also emerged as a key strength, with full agreement on the usefulness of the EPS, high compliance rates, and active participation of pharmacists in identifying gaps and proposing solutions. Another key strength of this study was the national site visit programme. Medical officers, pharmacists, and nurses from 28 hospitals across the country visited Hospital H-2, to observe the implementation of HAM safety practices. This programme provided participants with practical, hands-on exposure to standardized safety procedures across pharmacies, wards, and other clinical units. Following the demonstrations, many participants initiated similar practices in their own institutions. The initiative was facilitated and funded by the WHO in collaboration with the DHQS.

While the pilot implementation was successful, several study-related limitations were noted. The main limitations affecting scientific validity included the relatively small sample of hospitals and pharmacists, which may limit generalizability, and reliance on self-reported data, which could introduce reporting bias. Additionally, the short duration of the pilot may not have captured longer-term implementation challenges or sustainability of the EPS. Operational challenges such as limited space, inadequate technology (E.g. lack of colour printers, inadequate Wi-Fi, insufficient computer access), and the absence of strong error reporting systems may have affected the efficiency of implementation but do not impact the scientific validity of the study. Several barriers were identified applicable to healthcare staff and patients, underscoring the need for on-going education, infrastructure strengthening, and systematic monitoring.

In addition, involving only pharmacists for the study and not having nurses and doctors is also a major limitation as this study was focussed on the work of pharmacists. Nursing practices related to the administration of intravenous antibiotics require further reinforcement through targeted training and supervision. Furthermore, the study did not assess the direct impact of the intervention on the actual occurrence of HAM-related medication errors. As the study was conducted in a single tertiary care hospital, the generalizability of the findings may be limited, and there is a clear need for additional piloting in diverse healthcare settings, including peripheral hospitals and rural clinics.

The findings strongly support the nationwide scale-up of HAM safety practices across Sri Lanka. Key implications for policy and practice include the standardization of HAM operational procedures in all hospitals, strengthening of infrastructure such as storage facilities, counselling areas, and basic digital tools, and the integration of HAM safety modules into the education and training curricula of pharmacists, nurses and doctors. In addition, the introduction of periodic refresher training programmes and safety audits targeting the work of doctors and nurses in addition to pharmacists is recommended to sustain improvements over time related to usage of HAM. Enhancing patient education through multilingual resources and structured follow-up mechanisms will also be essential to reinforce safe medication practices and promote long-term patient engagement in medication safety.

## CONCLUSION

This study provides the first comprehensive evaluation of HAM and associated EPS in Sri Lanka. The HAM list was systematically developed based on Phase 1 studies and the ISMP list and validated through five consultative meetings with a large and diverse panel of medication safety experts and practicing pharmacists. Piloting its implementation in a tertiary hospital demonstrated the feasibility, acceptability, and high adherence of EPS among pharmacists, even in a resource-constrained environment. These findings highlight that structured, low-resource interventions can be successfully integrated into routine hospital workflows and have the potential to substantially reduce medication errors.

Importantly, this study directly informed national policy through the issuance of a MOH circular, demonstrating the real-world impact of evidence-based medication safety initiatives [27].

The results support the nationwide scale-up of HAM safety practices, including the integration of EPS into hospital operational procedures, reinforcement of staff training, and enhancement of patient education through multilingual resources. Future research should evaluate implementation across diverse healthcare settings, including peripheral and rural hospitals, involving all categories of healthcare professionals and explore the potential benefits of digital technologies to further reduce MEs. Overall, this study represents a significant step toward strengthening medication safety governance in Sri Lanka and provides a framework that can be adapted in other LMIC seeking to improve high-risk medication management.

## ACKNOWLEDGMENTS

Authors thankfully acknowledge the support of all the pharmacists and support staff of the Pharmacy Department of Colombo South Teaching Hospital - Kalubowila, The National Hospital of Sri Lanka - Colombo and pharmacists and pharmacy assistants of community pharmacies who participated in the study. Further thanks to staff of Patient Safety and Accreditation Bureau -PSAB (formerly DHQS-Directorate of Healthcare Quality and Safety) Ministry of Health.

## FINANCIAL DISCLOSURE

This project was not funded by any university research grant.

## COMPETING INTERESTS

The authors have declared that no competing interests exist.

## DATA AVAILABILITY STATEMENT

All relevant data are within the manuscript and its Supporting Information files. Qualitative data from focus group discussions contain potentially identifying information and cannot be shared publicly due to ethical restrictions. De-identified quantitative survey data are available upon reasonable request from the corresponding author.

## Supporting information

S1 Table. Total list of high-alert medications for acute care settings (total = 100)

S2 Table. Total list of high-alert medications for community settings (total = 40)

## Notes

### Competing Interest Statement

The authors have declared no competing interest.

### Author Declarations

Ethics approval and consent to participate was approved by the Ethics Review Committee of Faculty of Medicine, University of Colombo (Reference number: 18-008) and Ethical Review Committees of National Hospital of Sri Lanka (NHSL) (Reference number: AAj/ETH/COM/2017) and Colombo South Teaching Hospital (CSTH) (Reference number: PL/MO/2018-2019). All approvals were renewed annually and covered the full study period. Permissions from all selected community pharmacies were obtained prior to commencing the study. Participation was voluntary, and informed written and verbal consent were obtained from all participants.

